# Differences in predicted rates of vaginal births after cesarean across racial groups in a ‘race-neutral’ model

**DOI:** 10.1101/2023.10.14.23296978

**Authors:** Anjali Suresh, Katie O’nell

## Abstract

When physicians and pregnant patients make decisions about whether to pursue a vaginal birth or cesarean, there are many factors at play. While vaginal birth can have health benefits for both parent and child, there are significant safety risks. In order to minimize these risks, physicians use predictive models to determine how likely patients are to have successful vaginal births after cesareans (VBAC). For many years, these predictive models included race as a variable. This decision recently came under fire, and the Maternal Fetal Medicine Unit (MFMU) published a calculator that did not include race as a variable but still predicted VBAC success with high accuracy. A large body of work in machine learning has highlighted that supposedly de-biased systems often re-code sensitive variables like race in terms of proxy variables. In order to determine if this was the case in this calculator, we replicated their formula, then found base-rate statistics of all the input variables for three different racial groups: Black, White, and Asian. We found that the distribution of VBAC probabilities for our simulated patients from these three groups was indeed significantly different from each other. Further, the predicted VBAC rates increased as a function of societal marginalization: Black patients were 47.6% likely to have a successful VBAC, Asian patients had a 48.6% probability, and White patients had a 49.4% probability. While these values are all within a few percentage points of each other, the differences in these simulated distributions show how there may still be underlying disparities in the maternal healthcare system.

## Introduction

The use of technology in the medical industry has progressively increased in recent years as a way to provide quicker, more efficient, and patient-specific care. Although very beneficial, introducing these technologies into an industry that has a history of discriminatory notions against America’s marginalized groups could very easily impact the results of many of these advancements. The healthcare system has always been very apparent in how it is designed predominantly to benefit mostly white people while hurting and disregarding people of color. This underrepresentation has resulted in both pre-existing racial disparities in medicine and those exacerbated by medical technology. In skin disease diagnostics, for example, the diagnostic criteria are based on symptoms prevalent in white patients, resulting in underdiagnosis for patients with darker skin (Ly et al., 2021). This is made worse by technology when algorithmic diagnostic tools are fed only canonical examples of skin disease.

In this paper, we investigate the Maternal-Fetal Medicine Units Network’s (MFMU) vaginal birth after cesarean (VBAC) calculator, which predicts how likely a patient is to successfully give birth vaginally after a previous cesarean (Grobman et al., 2021). Initially, the MFMU calculator included a question that asked users whether they were African American and/or Hispanic. However, after the network recognized there was potential for racial bias that they wanted to address, they removed race and ethnicity as variables but still kept the accuracy rate of their predictions. So, we considered the fact that although efforts were made to eliminate any bias, the calculator could have still adopted proxy variables that were able to manipulate the results. This initiated our experiment, and we questioned whether we could develop a model of the calculator without requiring the use of race and ethnicity as variables and find possible explanations for where bias still arose. We started our experiment by replicating the VBAC calculator and then compiled a numerical dataset that included the averages of each variable: age, obstetric history, arrest of labor history, hypertension history, weight, and height for Black, White, and Asian women in America. We combined these two tools to create a patient simulator, which filled in the variables for the calculator based on whichever race was input. Then, we generated medical histories for 10,000 simulated patients and used the calculator to predict VBAC probability distributions and look for any differences in the probability likelihoods between each race.

## Background

Initially, our project’s scope was primarily on ethical computing and human rights; we then explored racism in the healthcare system before focusing on precision medicine and its flaws, which ultimately led to the investigation of the VBAC calculator. During our major focus on precision medicine, we examined three technical publications relating to the complexity of precision medicine in terms of how it handles race and ethnicity.

Long standing difficulties of prescribing medicine due to its impreciseness have made physicians consider implementing variables to better personalize care given to patients. So, while race is not a biological component, it correlates with geographic heritage, a driver of genetic variety that might alter drug response, and thus became a common variable in precision medicine. However, this only oversimplified the complexity of utilizing race in this manner (Bonham et al., 2016). Prescribing medications like this tends to overlook the importance of ancestry, health disease, and drug response. It suggested that using precision medicine to replace race as a medical variable will only work if the data being used is representative of the community that should benefit from it. This furthered the conversation on how the healthcare system lacks holistic representation in determining care for individuals. It also contributed to its claim by exhibiting different cases of precision medicine being faced with challenges pertaining to race and ethnicity to show us how a lot more needs to be done to perfect precision medicine before it can be used. For example, the first race-cased drug approved by the FDA, called BiDil, was used to treat heart failure in self-identified black patients. Some claimed that this was one step forward in revolutionizing personalized medicine, while others claimed that race as the only variable was not enough to be the determining factor of who gets to use the drug. This paper helped us in our research, as it gave insight into how precision medicine will never be a simple case and still needs a lot of work and significant change within the healthcare system to work.

In addition to understanding the ways that precision medicine exacerbates racial inequities, we wanted to learn more about how communities affected by racism perceive the use of predictive algorithms in medical care. A group of African American and Hispanic people were asked several questions pertaining to precision medicine to gather their perspectives on its benefits and disadvantages. The participants agreed that precision medicine is needed and could provide a lot of benefits, but were also scared that the potential disadvantages racism could present might cause more problems (Yeh et al., 2020). The paper helped us in our research as it gave us a viewpoint on direct feedback and perspectives on precision medicine from marginalized groups. But, because the participants came from different cities across the country, it’s unclear if the differences are because of cultural beliefs, historical and social situations, or a combination of the two. This paper lets us understand how there will always be a certain amount of distrust from these groups toward the medical industry due to the history they have with it.

From a broad perspective, it may appear that using factors such as race can perpetuate inequalities and reduce marginalized populations’ trust and buy-in, however, it can and does serve as a beneficial proxy when more granular data that would be useful is unavailable. Although it may seem unnecessary to use race/ethnicity as a variable in medicine, it could actually help capture trends for epidemiologic information. Race in the past has been used so much as a determiner for many factors today that there is no choice but to use it to look for these trends. In the past, racial injustice has compromised medicine through the exploitation of race and ethnicity as biological determinants, despite the reality that race and ethnicity have no biological basis. So, until better predictors are available, we should still use race/ethnicity to better address and eliminate health inequalities (Borrellet al., 2021). The paper contributed to our research as it also addressed the fact that the idea of race has become so complex and a major factor in our everyday lives that there is no point in being “color-blind”, but instead embracing it as a way to give us insight into other problems.

These papers were a crucial aspect of the start of our research, as it was necessary that we kept all of this information in the back of our heads throughout the process. We felt that it was necessary to keep an open mind throughout our research project in order not to undermine the complexity of using such a powerful variable as race in the healthcare system. Additionally, it motivated us to delve further into the potential sources of proxy variables and how they affect outcomes.

## Dataset

Our numerical dataset for this project consisted of a compilation of datasets that were pulled from seven different sources. We had 6 variables we needed to get race-based distributions for:

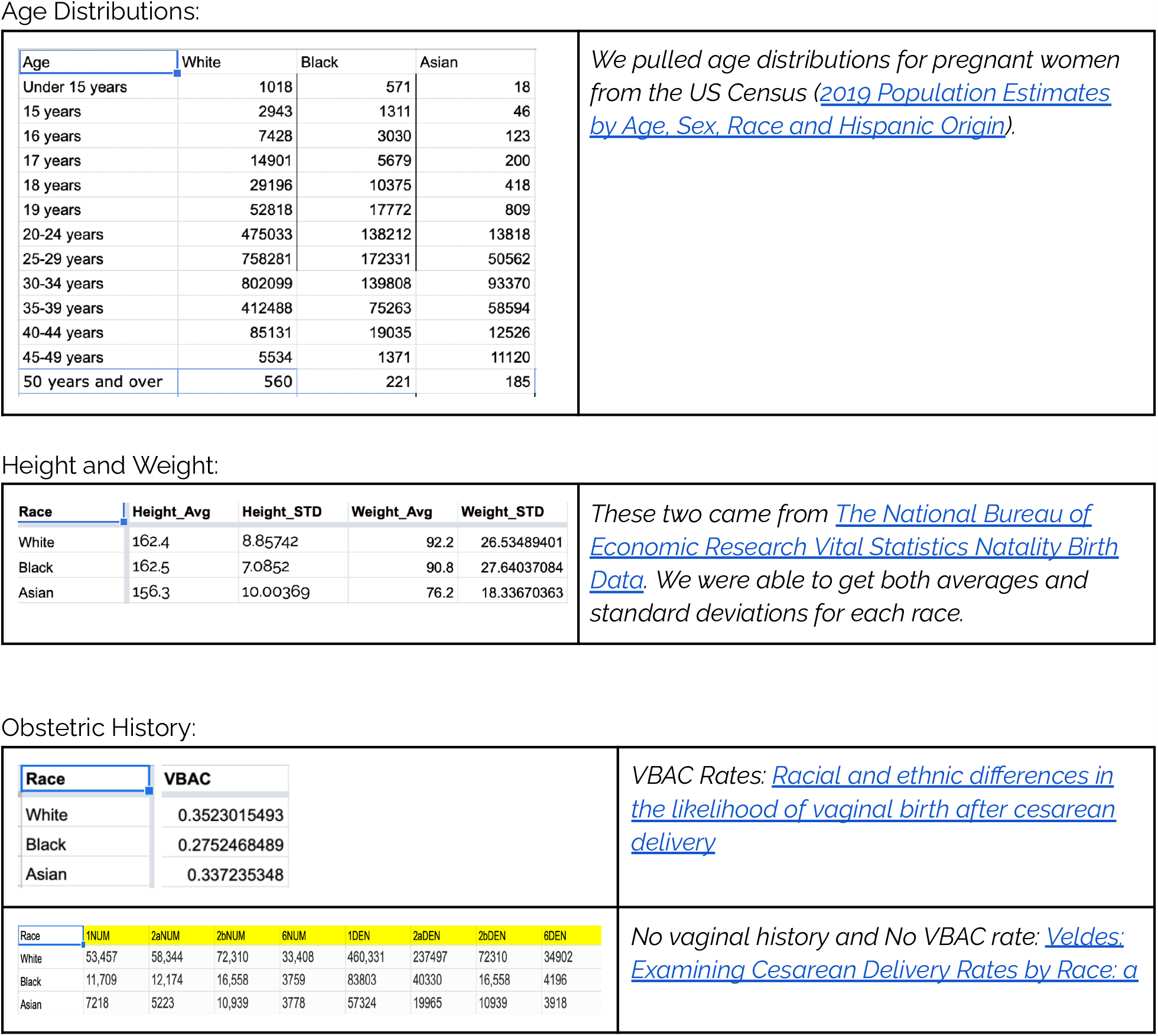

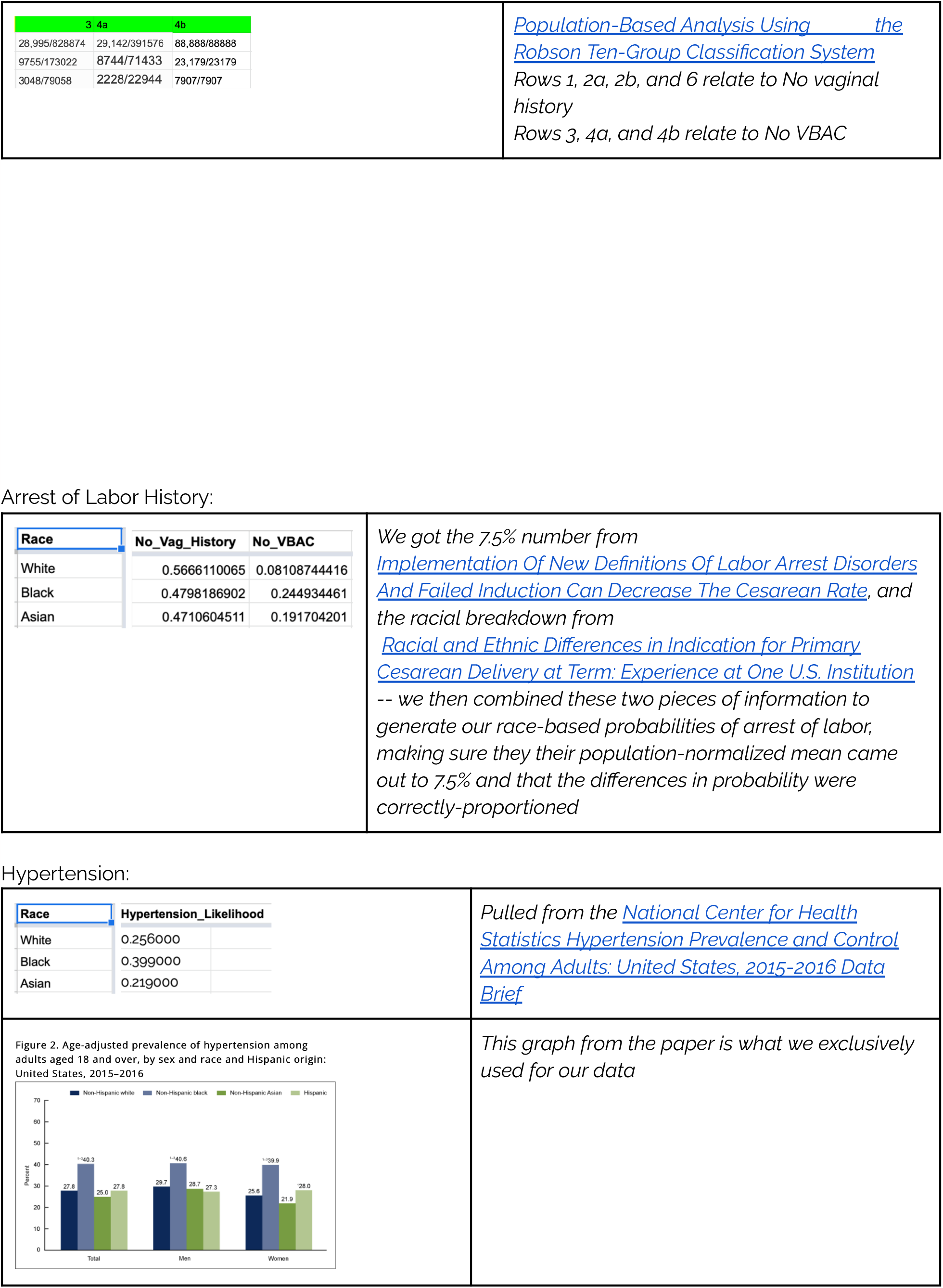

## Methodology and Models

Our main goal was to determine whether there is still encoded racial bias in the MFMU’s VBAC prediction calculator, even though race and ethnicity are no longer included. In order to answer this question, we looked to examine if the anticipated VBAC probabilities for Black, White, and Asian patients would differ systematically. First, we had to replicate the MFMU VBAC calculator using the paper’s linear model specified in their work (Grobman et al., 2021). Then, for each racial group, we identified the baseline distributions of the six input variables (age, height, weight, obstetric history, history of labor interruption, and hypertension). Using these distributions, we generated ten thousand simulated patients representing each of our three racial groups, forecast the likelihood that they would undergo a VBAC, and then compared the probability distributions to one another to check for any significant differences.

We were able to start the first step of our process of defining a replicate calculator in our code by using the weighted equation given by the MFMU network and naming it “predict_vbac”. Our calculator provides a probability score of the estimated likelihood of a vaginal birth following a cesarean depending on user inputs and has six parameters accounting for the six variables (questions) required by the calculator. Three of the variables require numbers as inputs within a defined range; these include: age (15-50 years old), height (119-191 cm), and pre-pregnancy weight (34-206 kg). Two of the variables are questions that require either a yes or no input; they are: arrest disorder indication for prior cesarean? and treated chronic hypertension? The last variable is for obstetrics history, which requires one of three possible inputs: previous VBAC, no previous vaginal history, or previous vaginal delivery only before a prior cesarean. After creating our calculator, we tested its accuracy compared to the MFMU calculator in three different test cases by seeing the difference between both probability scores after entering the same random inputs for each variable. On average, our calculator’s scores were off by only 0.5% of the MFMU’s calculator for all three test cases.

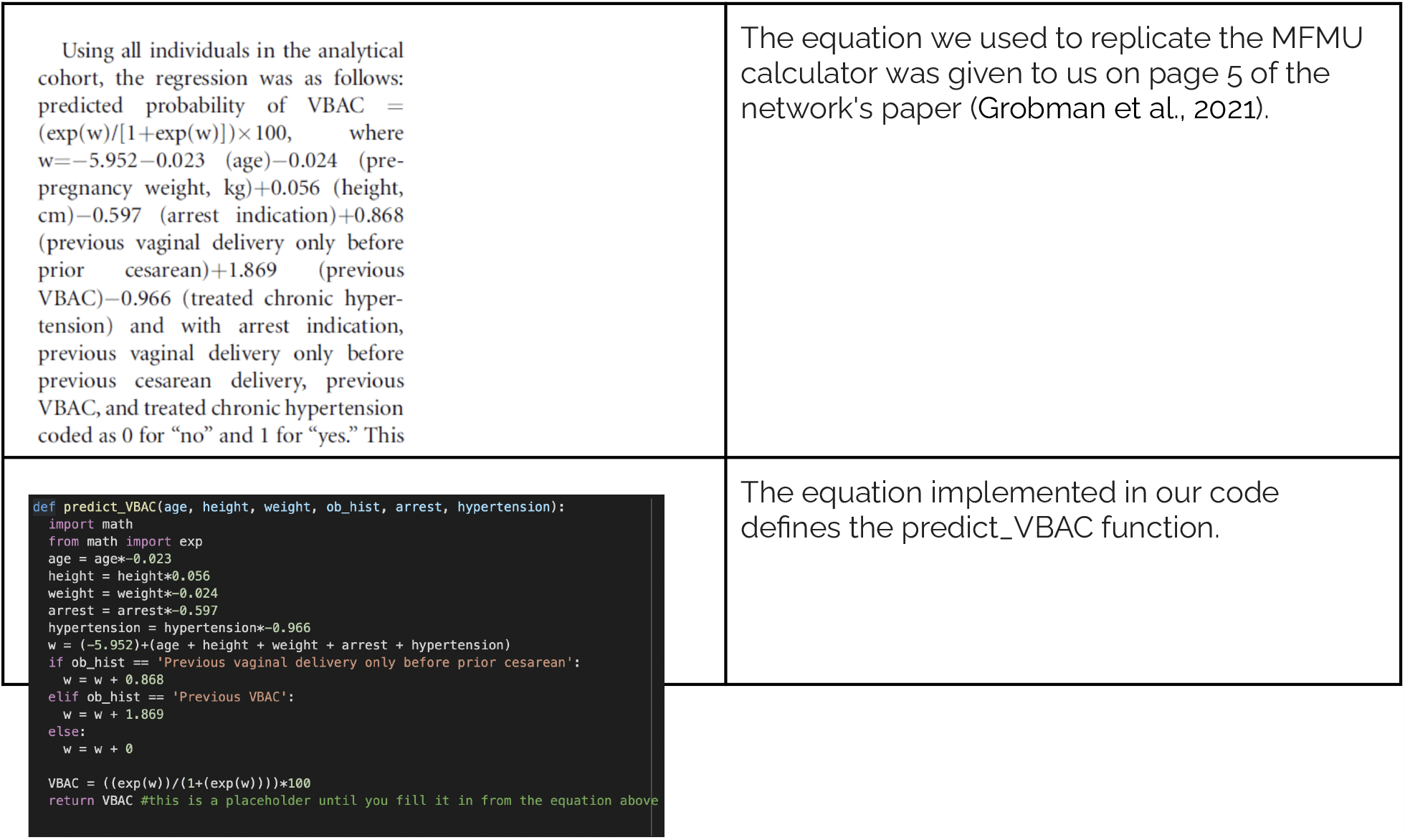

The next step in our process was to create our simulated patient generator, which we called “get_simulated_patient”. We had to first compile a large dataset with data derived from multiple sources, which contained: the average heights and their standard deviation, average weights and their standard deviation, arrest likelihoods, hypertension likelihoods, age distribution for pregnancies, and averages of the three inputs for OB history; the variables were each separated by race. We then imported this data back into our code. Using this, we were able to create a randomized representative sample for each of the variables based on the three races. Height and weight were both safely assumed to have a normal distribution, as they were the only ones that contained averages along with standard deviations, so we used the “numpy.random.normal()” function to get our representative samples. Arrest likelihoods, hypertension likelihoods, and OB history were categorical variables determined by the probability of occurrence. For these variables, we used the function “numpy.random.random()”, which works essentially like a weighted coin; the function generates a random number between 0 and 1, and if that number lands below the likelihood of that event happening, the function is true. Finally, for age distribution, the data we were using was already a representative sample, so we implemented the “numpy.cumsum()” function and used the “numpy.random.random()” function once again to generate a random age. The “get_simulated_patient” function returns a randomized representative sample for all six variables.

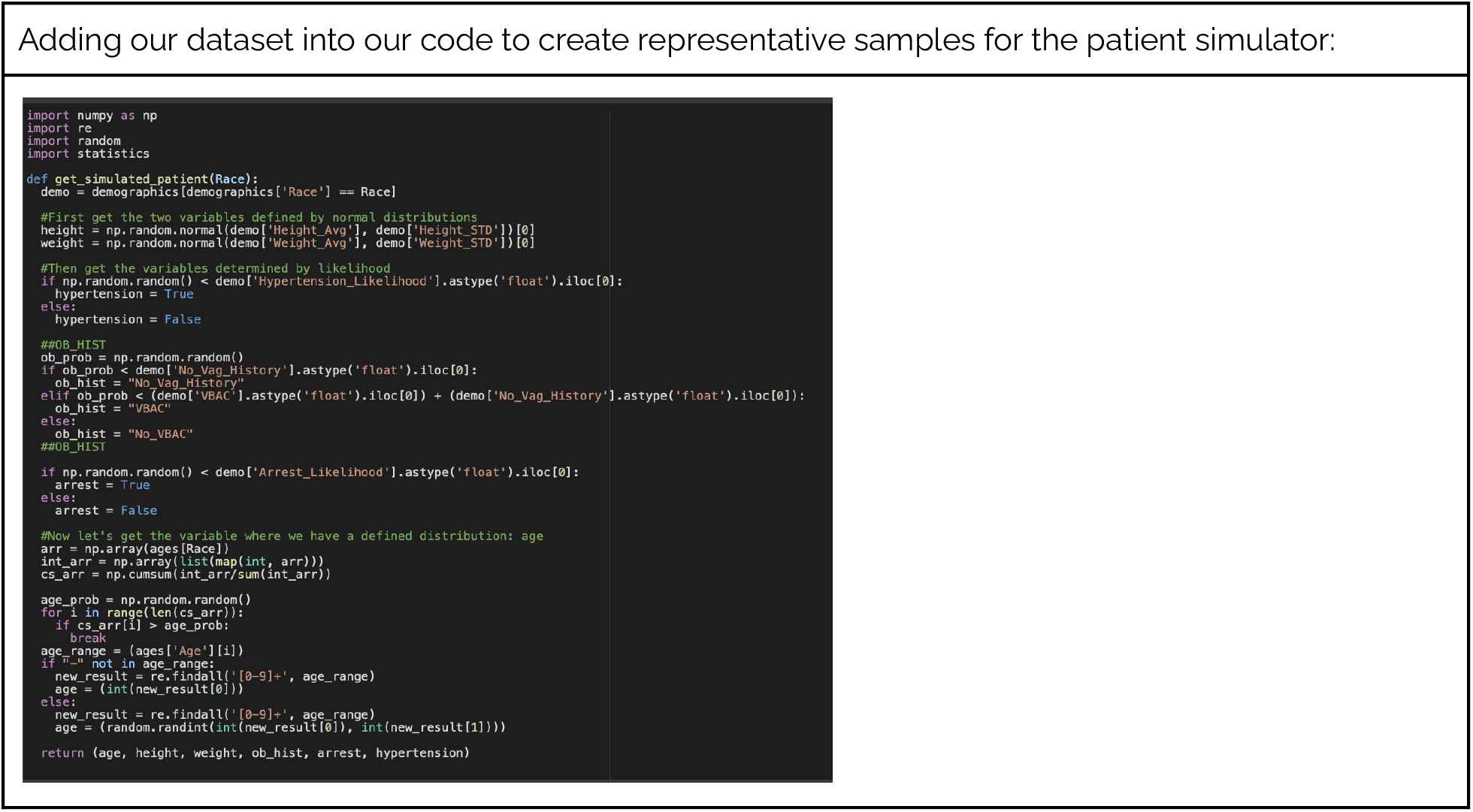

For our last step, we used the “get_simulated_patient” along with the “predict_VBAC” function to create 10,000 randomized patient histories for each race and output their VBAC scores. Then, we used Matplotlib’s plt.hist() function to plot a histogram for all the scores. This was done to check for any differences in the overall distribution of the graphs and their averages. Then we conducted a t-test for independence and checked if there were any significant differences among the mean probabilities.

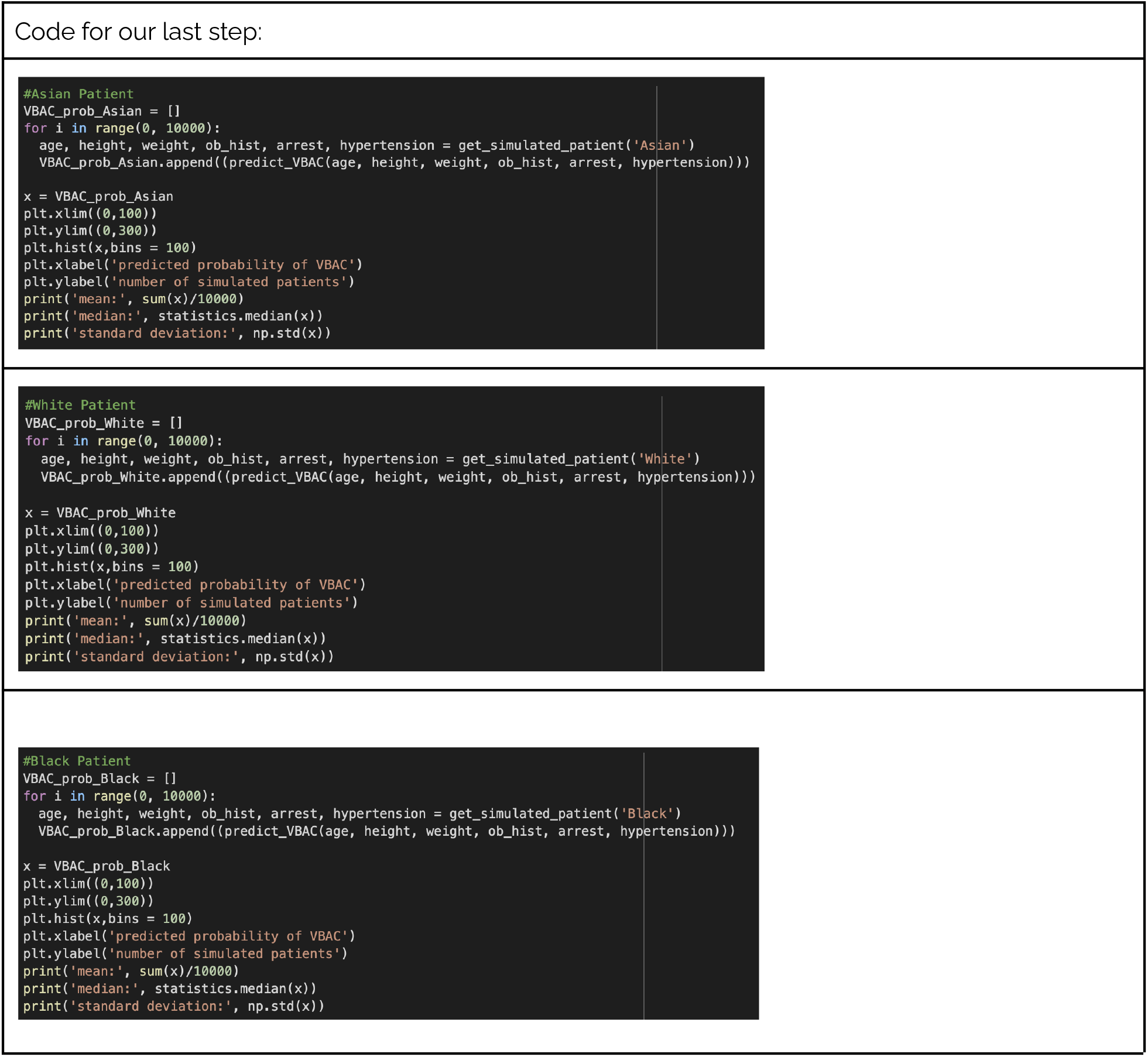

## Results and Discussion

By eye, the distributions of VBAC probabilities for Asian, Black, and White patients look quite similar. However, we later conducted three pairwise t-tests comparing each patient group to each other. After Bonferonni correction for multiple hypothesis testing, the distributions of Black-White and Black-Asian VBAC scores were significantly different from each other (p-values of 4e-14 and 9e-09 respectively). The differences in the means between each of the populations were minimal, staying within the 1-2% range. The fact that the distributions of predicted VBAC scores were different for different racial groups suggests that the original calculator might have re-coded race in terms of other variables. Moving forward, it would likely be useful to build a linear model that predicts a patient’s race from their biological variables for our simulated patients in order to determine which biological variables are most likely to be a proxy for race.

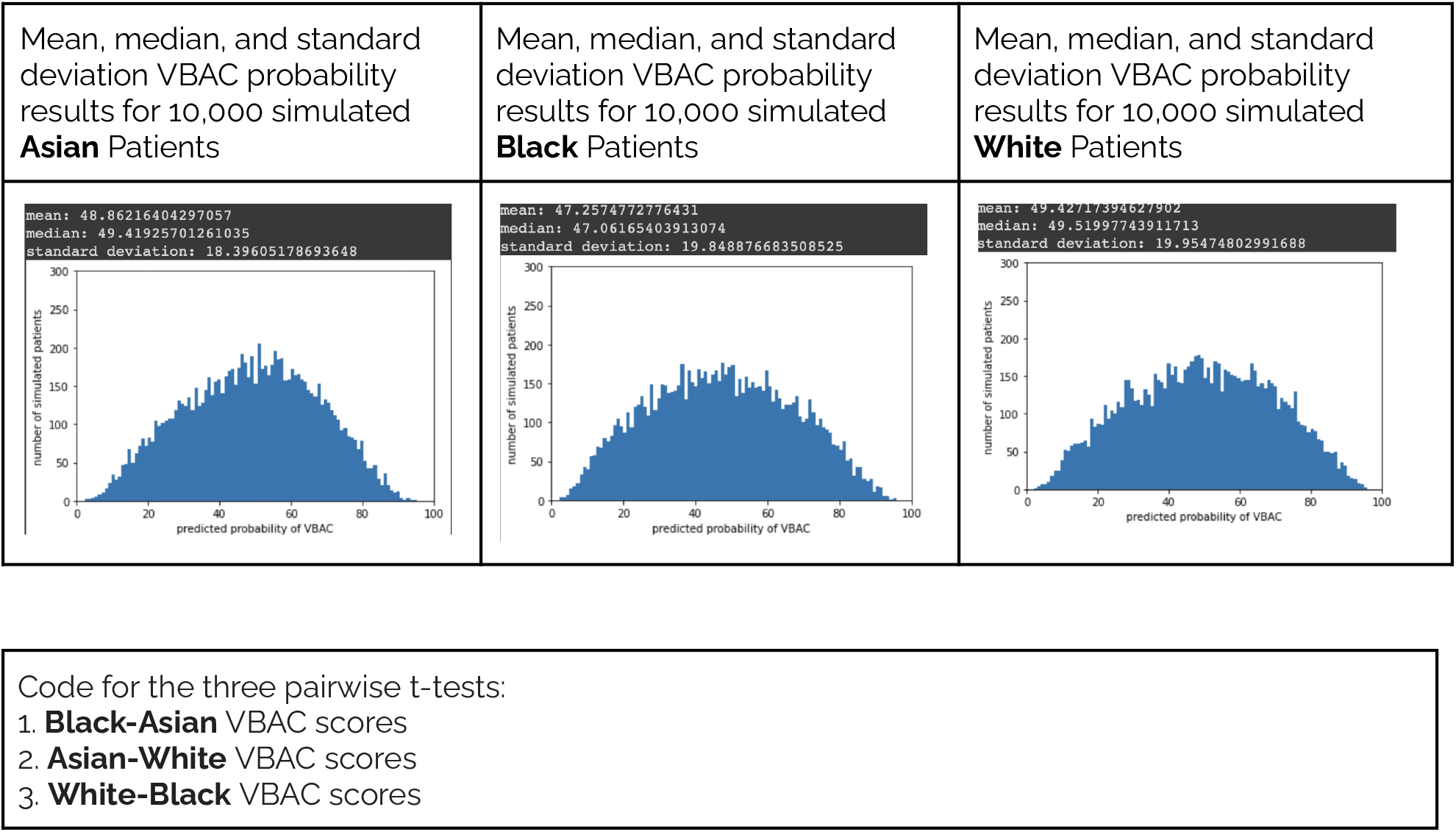

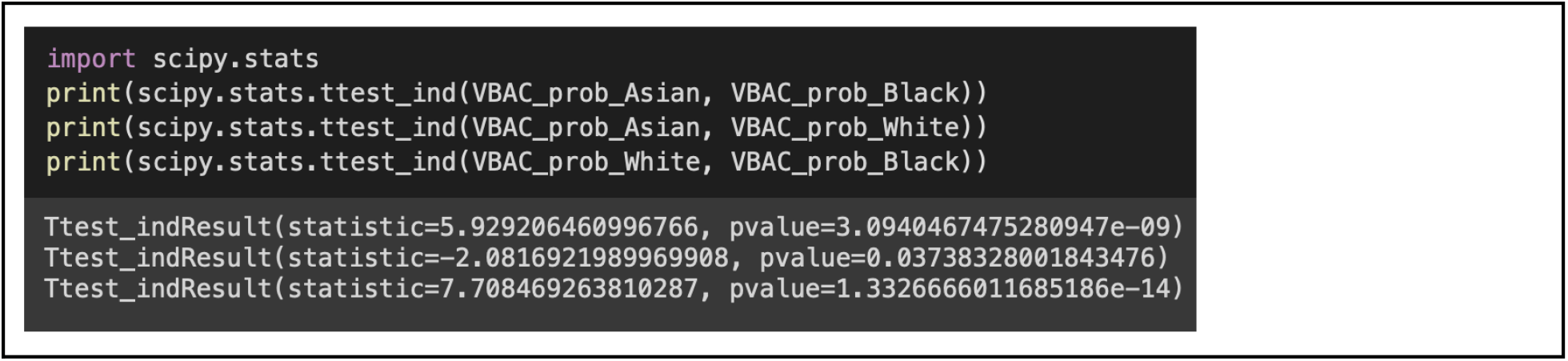

## Conclusion

The main motivation of our research was to understand the impact of proxy variables hidden within the VBAC calculator. The results from our t-test tell us that the difference was not due to chance or a sampling error, but that there still remain underlying factors that are the cause of these 1-2% differences in the averages. These factors can range from underrepresentative samples to the overall healthcare given to black people in America. The next steps that should be taken with this research are to find datasets that are more representative of all three races, including ethnicity, and to determine which biological variables proxy race. In consideration of the overall usage of similar algorithms in the healthcare system, it would be beneficial if we began to consider ethical computing as a prerequisite to the design processes of these advancements and were able to identify how and why bias can infiltrate into the makeup of these algorithms. We can also attempt to dive further into a more non-medical and public health setting to understand how the social determinants of health (SDOH) have been able to impact such variables. Implementing such research before creating medical technology should be done as a way to educate ourselves about our community and understand how marginalized groups are disproportionately disadvantageous in almost every aspect of the SDOH.

## Data Availability

All data produced in the present work are contained in the manuscript

https://www.census.gov/newsroom/press-kits/2020/population-estimates-detailed.html

https://www.nber.org/research/data/vital-statistics-natality-birth-data

https://www.ncbi.nlm.nih.gov/pmc/articles/PMC4618667/

https://link.springer.com/article/10.1007/s40615-020-00842-3/tables/2

https://elischolar.library.yale.edu/cgi/viewcontent.cgi?article=3401&context=ymtdl

https://www.researchgate.net/publication/256448154_Racial_and_ethnic_differences_in_primary_unscheduled_cesarean_deliveries_among_low-risk_primiparous_women_at_an_academic_medical_center_A_retrospective_cohort_study

https://www.cdc.gov/nchs/products/databriefs/db289.htm

